# Environment-Wide Association Studies of Anemia in the National Health and Nutrition Examination Surveys

**DOI:** 10.1101/2023.06.02.23290861

**Authors:** Jiayan Zhou, Nicole Palmiero, Kristin Passero, John R McGuigan, Tomás González Zarzar, K. Sandeep Prabhu, Molly A. Hall

## Abstract

**Background:** Anemia is a global health problem that can lead to chronic illness in adults and may be fatal in children and the elderly. While some dietary factors and heavy metals are known risk factors for anemia, there are no environment-wide studies of anemia.

**Objectives:** Our goal was to identify environment-wide risk factors for anemia.

**Methods:** We evaluated general anemia in children and adults and further classified anemia as a) iron, vitamin B12, or folate deficiency anemia; b) anemia in general chronic diseases; and c) anemia in chronic kidney disease. As well as quantitative measures including level of hemoglobin, serum vitamin B12, red blood cell (RBC) folate, and serum iron. Environment-wide association studies (EWAS) were performed to identify novel environmental risk factors of anemia in discovery and replication subsets of the National Health and Nutrition Examination Surveys (NHANES).

**Results:** We identified and replicated 106 potential environmental risk factors for anemia. As expected, serum iron was the top exposure associated with general anemia for adults. Cadmium was associated with adult hemoglobin levels, as were vitamin Bs, micronutrients, smoking, and alcohol consumption. Further, decreased levels of multiple vitamins, including vitamin A, vitamin Es and multiple vitamin Bs, were associated with general anemia in adults. Use of tobacco and alcohol was also found to be associated with red blood cell folate and serum iron levels. In children, serum iron level was associated with folic acid supplements and vitamin A supplements.

**Discussion:** This is the first EWAS of anemia, providing insights into the environmental etiology of anemia risk in children and adults. These results may lead to the development of public health recommendations to mitigate anemia risk factors.

## Introduction

Because hemoglobin in red blood cells (RBC) carries oxygen, reducing either decreases oxygen availability, leading to anemia and fatigue, especially in young children and pregnant women (de Benoist et al. 2005). In the US, anemia and iron deficiency led to 710000 emergency department visits in 2018 (Center for Health Statistics 2018) and 5563 deaths in 2020 (Centers for Disease Control and Prevention 2022). In the US in 2019, the prevalence of anemia in women of reproductive age (aged 15–49) was 11.8% (WHO 2021), and in young children (aged 6–59 months) was 6.1% (WHO 2021a).

A diagnosis of anemia is based on RBC counts, hemoglobin (de Benoist et al. 2005), mean corpuscular volume (MCV) (Maner and Moosavi 2019), and abnormal blood cell morphologies (Losek et al. 1992), even without clinical signs and symptoms (American Society of Hematology). Anemia’s risk factors include bleeding, infection, inflammation, nutritional deficiencies, and poisons (Shamah et al. 2016). Anemia can be classified into different types, including nutritional deficiency anemia (iron, vitamin B12, or folate deficiencies), anemia in general chronic diseases, and anemia in chronic kidney disease (de Benoist et al. 2005). Iron deficiency anemia (IDA), the most common anemia in developing and developed countries, may result from blood loss, low iron consumption or absorption, gastrointestinal surgery, or kidney failure (National Heart Lung and Blood Institute 2019). The standard treatment for IDA is iron supplementation with screening for inflammatory conditions and diseases (National Heart Lung and Blood Institute 2019). However, environmental factors also affect IDA. For example, lead in water promotes pediatric IDA, while vitamin C enhances iron absorption, thereby reducing IDA. Vitamin B12, which is derived from animal and dairy products, is also essential for producing healthy RBCs (National Heart Lung and Blood Institute). A deficiency in vitamin B12 can reduce RBCs, promote abnormal iron absorption, and lead to anemia due to dysfunction of the stomach or small intestines. Air pollution such as PM_2.5_ particles and NO_2_ are also associated with anemia in chronic diseases (Honda et al. 2017).

Environmental exposure impacts many complex diseases, and environmental factors including diet, supplements, and pollution contribute to anemia. Environment-wide association studies (EWAS) use high-throughput techniques to identify associations between environmental factors and complex traits such as body mass index (Lucas et al. 2019) or diseases, including type II diabetes (Hall et al. 2014; Patel et al. 2010), chronic kidney disease (CKD) (Lee et al. 2020), left ventricular diastolic dysfunction (Dhall et al. 2021), and peripheral arterial disease (Zhuang et al. 2018). However, EWAS have not been used to characterize the environment-wide risk factors for anemia. Here, we performed EWAS to identify potential environmental risk factors for different types of anemia using more than 40,000 participants from four cohorts in the NHANES and 963 environmental exposures.

## Materials and Methods

### National Health and Nutrition Examination Survey (NHANES)

The NHANES are conducted by the Centers for Disease Control and Prevention (CDC) to evaluate the health and nutritional status of the US population (Cousins 2014; Le 2016; McFarlane et al. 2008; Patel et al. 2009). The goal of NHANES is to identify risk factors for prevalent diseases to develop effective public health policies. The participants’ demographic information, dietary recalls, health surveys, toxin exposures, and laboratory measurements were collected by interviews and physical examinations at home or a mobile exam center (Cousins 2014). The current study, including 41,474 individuals and 1,191 variables before quality control (QC) from survey years 1999–2006, was pulled from the unified dataset (Patel et al. 2016). We downloaded data for pregnancy status, RBC folate measurement (nmol/L), serum vitamin B12 measurement (pmol/L), frozen serum iron measurement (μmol/L), refrigerated serum iron measurement (μmol/L), and erythrocyte protoporphyrin measurement (μmol/L) as .XPT files from the NHANES website for each survey cycle (https://www.n.cdc.gov/nchs/nhanes/default.aspx). The .XPT files were imported into R software with the implemented package called SASxport. Because of changes in the technology to measure ferritin concentration levels in the 2003−2006 survey cycles, a correction to the 1999−2002 concentration values (ng/mL) was applied as follows (Miller 2014):

If ferritin (ng/mL) ≤ 25, then corrected ferritin (ng/mL) = 1.2534 × ferritin (ng/mL) + 1.4683; if 25 < ferritin (ng/mL) ≤ 65, then corrected ferritin (ng/mL) = 1.2001 × ferritin (ng/mL) + 1.4693; or if ferritin (ng/mL) > 65, then corrected ferritin (ng/mL) = 1.0791 × ferritin (ng/mL) + 4.8183.

### Anemia Classification

General anemia cases were diagnosed using the World Health Organization (WHO) criteria (WHO 2011) using hemoglobin threshold levels based on age, sex, and pregnancy status (Table 1). Anemia cases were further classified as nutritional deficiency anemia (IDA, vitamin B12 deficiency anemia, and folate deficiency anemia), chronic kidney disease-related anemia (CKD-A), chronic disease-related anemia (CDA), and unexplained anemia (UA) (Figure 1 and Table 2). IDA was defined by the presence of at least two of three standards: 1) transferrin saturation < 15%, 2) serum ferritin < 12 ng/mL, or 3) erythrocyte protoporphyrin > 1.24 μmol/L (Guralnik et al. 2004; Knovich et al. 2008; Parikh et al. 2011; Patel et al. 2009). Vitamin B12 deficiency anemia was defined as serum vitamin B12 < 147.56 pmol/L (Guralnik et al. 2004; Knovich et al. 2008; Patel et al. 2009). Folate deficiency anemia was defined as RBC folate < 232.49 nmol/L at a mobile examination center (Guralnik et al. 2004; Patel et al. 2009). CKD-A was defined as glomerular filtration rate (GFR) < 60 mL/min/1.73m^2^ for the cases without nutrient deficiency anemia (Astor et al. 2002; Coresh et al. 2007; Weiss et al. 2019a). CDA was defined as serum iron < 10.74 nmol/L for individuals with total body iron (TBI) < 0 mg/kg for the cases without nutrient deficiency-induced anemia and CKD-A (Gupta et al. 2017; Guralnik et al. 2004; Park and Eicher-Miller 2014; Patel et al. 2009). UA included individuals with anemia that could not be classified into another subtype (Guralnik et al. 2004; Patel et al. 2009). The GFR was calculated for participants based on their serum creatinine (mg/dL), age in years, sex, and race/ethnicity by equation 1 (Coresh et al. 2007). The TBI was determined based on Flowers soluble transferrin receptor (sTfR) by equation 2, and ferritin concentration by equation 3 (Cogswell et al. 2009; Gupta et al. 2017).

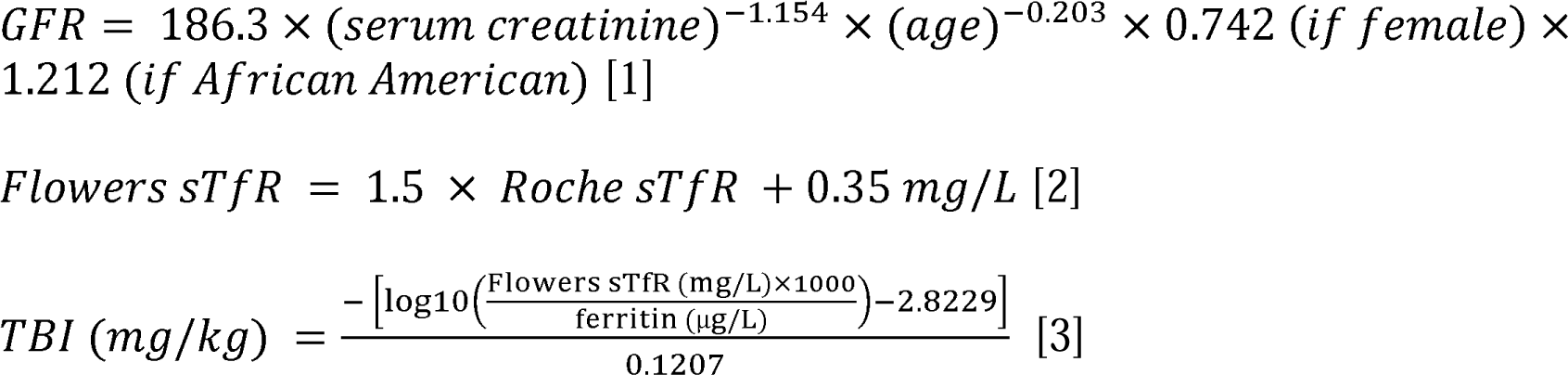

**Table 1.** Anemia screening based on hemoglobin thresholds from WHO.

| Group | Age in years | Sex | Hemoglobin (g/dL) |
| --- | --- | --- | --- |
| Children | 0–4.99 | Male and female | ≤ 11.0 |
|  | 5–11.99 | Male and female | ≤ 11.5 |
|  | 12–14.99 | Male and female | ≤ 12.0 |
| Adults | ≥ 15 | Male | ≤ 13.0 |
|  |  | Pregnant female | ≤ 11.0 |
|  |  | Nonpregnant female | ≤ 12.0 |

Notes: Anemia cases are defined based on hemoglobin levels from the WHO standards with age in years, sex, and pregnancy status for a female more than 15 years old (WHO 2011).

**Figure 1.**
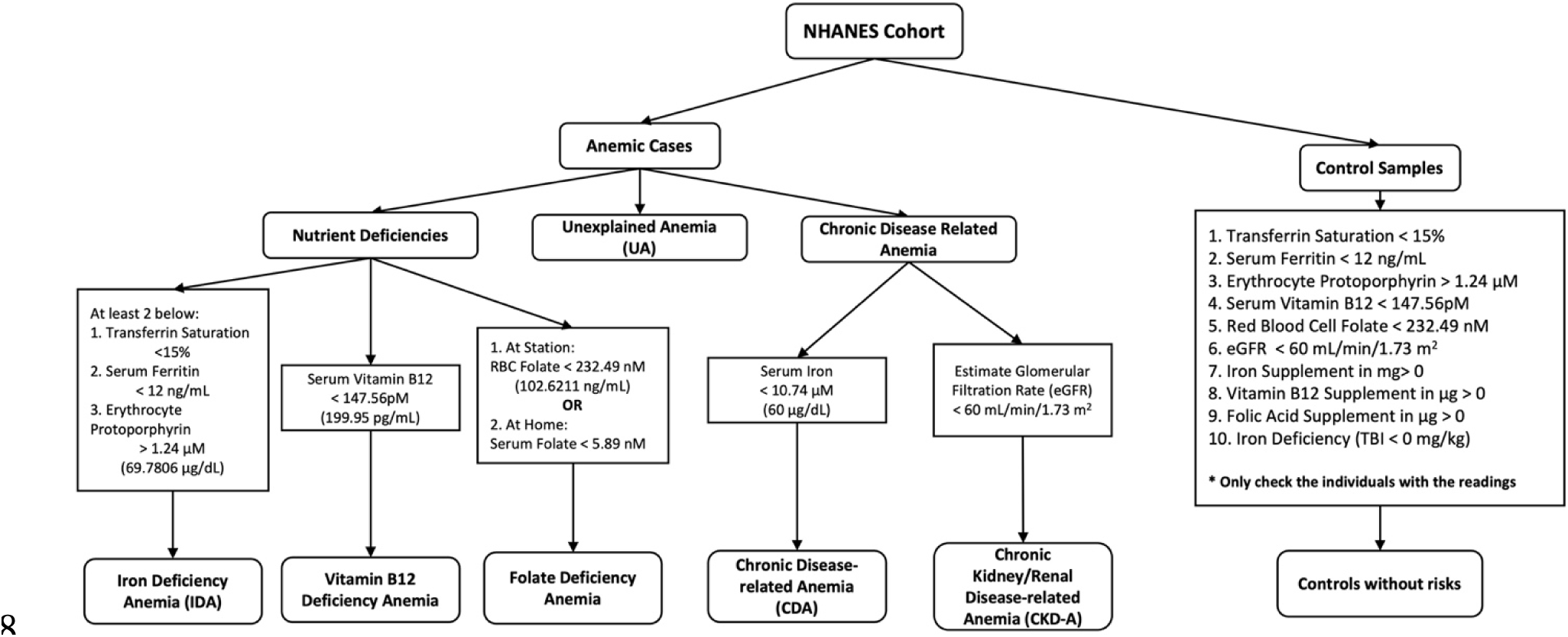
Determination and classification of anemic and control participants. Individuals without hemoglobin measurements were eliminated from the study. Females without a pregnancy test, missing a pregnancy test value, or with an invalid test were considered nonpregnant. The anemia cases were based on the WHO standards (WHO 2011). The anemic participants were classified into five subgroups shown at the bottom of the panel. Individuals with pre-anemic symptoms were eliminated from the control group. Only individuals with clinical test values were included. Individuals with missing values in the standards were considered to be removed from the control participants.

**Table 2.** Summary of the sample sizes for anemic and non-anemic individuals in NHANES.

| **Population** | **1999–2000** | **2001–2002** | **2003–2004** | **2005–2006** | **Total** |
| --- | --- | --- | --- | --- | --- |
| General anemia | 517 | 610 | 527 | 559 | 2213 |
| Iron deficiency anemia | 177 | 247 | 127 | 162 | 713 |
| Vitamin B12 deficiency anemia | 15 | 23 | 12 | 18 | 58 |
| Folate deficiency anemia | 6 | 3 | 5 | 2 | 16 |
| Chronic disease-related anemia | 0 | 0 | 35 | 34 | 69 |
| Chronic kidney disease-related anemia | 45 | 53 | 79 | 95 | 272 |
| Unexplained anemia | 282 | 299 | 274 | 258 | 1113 |
| No anemia | 4165 | 4068 | 4263 | 3949 | 16445 |

### Exclusion Criteria for Control Samples

People who had normal hemoglobin levels but met one or more criteria for any anemia subtype were classified as pre-anemic. Individuals with pre-anemia symptoms or risk factors for developing anemia, such as low transferrin saturation, low serum ferritin concentration, and high erythrocyte protoporphyrin level, were excluded from the study (Figure 1 and Table 2). Participants taking iron, vitamin B12, or folic acid supplements were also excluded from the study, as these individuals could potentially be treating their anemia with supplements or taking preventative measures against anemia. Participants who were not in the anemia group and were not excluded from any criteria established previously were used as controls.

### Study Design

We employed a discovery and replication study design, whereby EWAS were performed in a discovery subset of NHANES (survey cycles in 1999−2000 and 2001−2002) and validated in a replication subset (survey cycles in 2003−2004 and 2005−2006) (Table 3).

**Table 3.** Participants by cohort, age, and ethnicity for NHANES

| Study | Survey years | Study cohort | Number of people | age < 15 years | female | Caucasian American | African American | Mexican American | Other Hispanic | Other ethnicity |
| --- | --- | --- | --- | --- | --- | --- | --- | --- | --- | --- |
| NHANES | 1999-2002 | Discovery | 21004 | 36.8 % | 51.4% | 38.0% | 23.4% | 29.4% | 5.2% | 4.0% |
|  | 2003-2006 | Replication | 20470 | 36.9 % | 50.9% | 39.4% | 26.2% | 26.2% | 3.4% | 4.8% |

Note: The number of individuals was reported for each study with a breakdown by cohort, age, and ethnicity. NHANES: National Health and Nutrition Examination Survey.

Four case-control EWAS and four EWAS with quantitative outcomes were performed (Table 4). The case-control analysis included general anemia, IDA, CKD-A, and UA. The IDA case-control study design was restricted to females, as males with IDA were not available in the replication dataset. Analyses for the remaining anemia subtypes, including vitamin B12 deficiency anemia, folate deficiency anemia, and CDA were not performed due to small sample sizes. Therefore, we used vitamin B12, RBC folate, and iron serum concentration levels to explore potential intermediate phenotypes in anemia. The fourth quantitative study design used hemoglobin serum levels to identify other potential risk factors in anemia.

**Table 4.** Study designs for EWAS.

| Types of study | Analysis | Population | Covariates |
| --- | --- | --- | --- |
| Case-control EWAS | General anemia vs. no anemia | Children | Age, sex, race, socioeconomic status, and survey year |
|  |  | Adults | Age, sex, race, socioeconomic status, survey year, and cotinine |
|  | Iron deficiency anemia vs. no anemia | Female adults only | Age, sex, race, socioeconomic status, survey year, cotinine, and pregnancy status |
|  | Chronic kidney disease-related anemia vs. no anemia | Adults | Age, sex, race, socioeconomic status, survey year, and cotinine |
|  | Unexplained anemia vs. no anemia | Children | Age, sex, race, socioeconomic status, and survey year |
|  |  | Adults | Age, sex, race, socioeconomic status, survey year, and cotinine |
| Continuous EWAS | Anemia - hemoglobin | Children | Age, sex, race, socioeconomic status, survey year, cotinine, and iron supplement intake |
|  |  | Adults |  |
|  | Vitamin B12 deficiency anemia - serum Vitamin B12 | Children | Age, sex, race, socioeconomic status, and survey year, dietary VB12, and VB12 supplementation |
|  |  | Adults |  |
|  | Folate deficiency anemia - RBC folate | Children | Age, sex, race, socioeconomic status, survey year, and folic acid supplementation |
|  |  | Adults |  |
|  | Iron deficiency and chronic disease-related anemia - serum iron | Children | Age, sex, race, socioeconomic status, and survey year |
|  |  | Adults |  |

Note: There are four case-control studies and four individual quantitative outcome approaches. For covariates, race categories are African American (NHANES label: black), Caucasian American (white), Mexican American, other Hispanic (other\_hispanic), and other ethnicities (other\_eth). Sex categories are female and male. Survey years are labeled based on a two-year cycle, for example, individuals surveyed in the years 1999 and 2000 were classified in one survey year cycle (Survey year = 1). Other covariates are continuous variables. EWAS: Environment-wide association study.

Following the WHO standards for anemia, quality control (QC) protocols and analyses were performed separately for children (age < 15 years) and adults (age ≥ 15 years) (WHO 2011). Therefore, each of the eight EWAS was divided into adults and children when possible. Replication of CKD-A in adults and UA in children was precluded by the small sample sizes (less than 200 samples). Instead of dropping these two populations from the survey cycles in 2003−2004 and 2005−2006, we merged the discovery and replication subsets into a single cohort to identify associated risk factors. We also performed a study using frozen serum iron levels and the combined serum iron from frozen and refrigerated samples to identify children with iron deficiency and/or CDA to access consistency of because both methods were used to store the individual blood samples. The combined serum iron from frozen and refrigerated samples was expected to provide more robust results because the number of frozen serum samples was limited.

### Quality Control

The QC steps for each study design were performed separately using CLARITE (Lucas et al. 2019; Passero et al. 2020; Zhou et al. 2020) with 963 environmental variables. For QC, 1) individuals with missing values for the phenotype or covariates were excluded; 2) variables were classified into binary, categorical, and continuous; 3) histograms were plotted for visualizing the distribution of the quantitative phenotypes and a log(x + 1) transformation was performed to normalize the data and reduce the high skewness for serum vitamin B12, serum folate, and serum iron; 4) variables for which 90% or more of the samples had values of zero were removed; 5) variables with n < 200 were dropped, and categorical/binary variables with a category size < 150 were removed (Table 5 and Table 6). Finally, only variables with the survey weights present in both discovery and replication subsets for cases and controls were considered in the study (Variable Dictionary: Excel Table S1).

**Table 5.** The number of participants before and after quality controls.

| Approaches | Study | Population | Pre-quality control |  | Input in EWAS after QC and cross-checking |  |
| --- | --- | --- | --- | --- | --- | --- |
|  |  |  | Discovery | Replication | Discovery | Replication |
| Case-control approach | General anemia | Adults | 903 | 892 | 712 | 794 |
|  |  | Children | 224 | 194 | 173 | 164 |
|  | Control | Adults | 5859 | 5133 | 5019 | 4787 |
|  |  | Children | 2374 | 3079 | 2143 | 2647 |
|  | IDA | Adults | 50 males + 316 females | 0 males + 260 females | 262 females | 241 females |
|  | CKD-A | Adults | 272 | - | 164 | - |
|  | UA | Adults | 417 | 388 | 330 | 339 |
|  |  | Children | 308 | - | 245 | - |
| Quantitative outcome approach | Hemoglobin | Adults | 26179 |  | 9744 | 10612 |
|  |  | Children | 15295 |  | 3810 | 3888 |
|  | Serum VB12 | Adults | 26179 |  | 9579 | 10410 |
|  |  | Children | 15295 |  | 3749 | 4038 |
|  | RBC folate | Adults | 26179 |  | 9986 | 10608 |
|  |  | Children | 15295 |  | 3991 | 4429 |
|  | Serum iron | Adults | 26179 |  | 10021 | 10693 |
|  |  | Children | 15295 |  | 4289 | 1561 |
|  |  | Children(c) | 15295 |  | 4293 | 1561 |

Note: The number of individuals was reported for each study in each step. A cross-check was performed, and the variables that are available with the survey weights in both the discovery and replication datasets for cases and controls were considered for running EWAS. QC: quality control. IDA: iron deficiency anemia. CKD-A: chronic kidney disease-related anemia. UA: unexplained anemia. EWAS: environment-wide association study. Children(c): combined readings of the blood iron (µg/dL) from frozen serum and refrigerated serum.

**Table 6.**
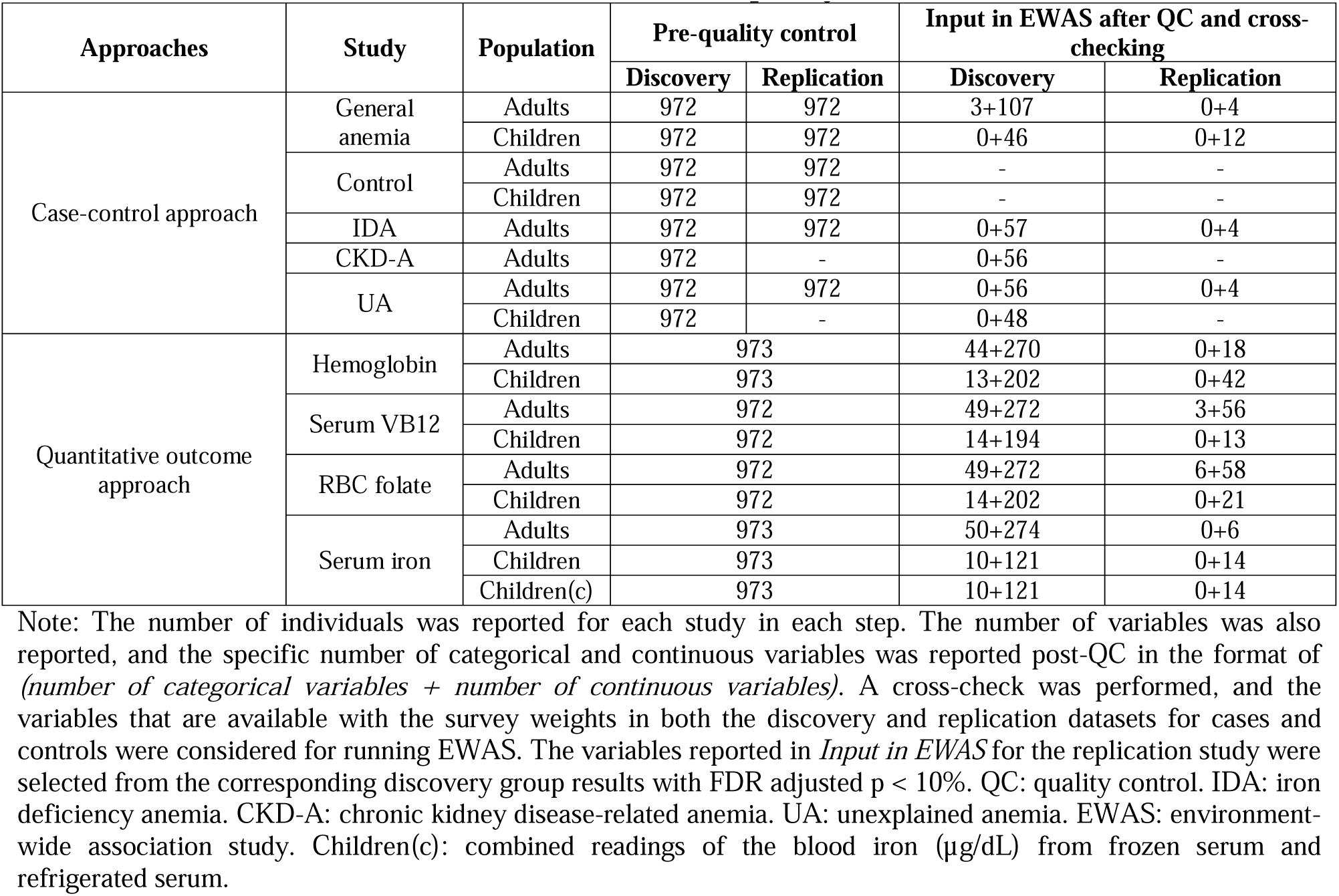
The number of variables before and after quality control

The numbers of variables remaining following QC are summarized in Table 6. Summary statistics and demographic information for participants are reported in Table 7, including the number or percentage of people in each cohort (discovery cohort), sex (female), age group (age < 15 years), and ethnicity (European American, African American, Mexican American, other Hispanic, and other ethnicities).

**Table 7.** Participants by cohort, age, and ethnicity after quality control

| Approaches | Study | Case/Control | Input in EWAS after QC and cross-checking |  |  |  |  |  |  |  |  |
| --- | --- | --- | --- | --- | --- | --- | --- | --- | --- | --- | --- |
|  |  |  | Number of people | Number of people from discovery cohort | age < 15 years | female | Caucasian American | African American | Mexican American | Other Hispanic | Other ethnicity |
| Case-control approach | General anemia | Case | 1843 | 885 | 337 | 1292 | 25.5% | 51.4% | 16.0% | 3.7% | 3.4% |
|  |  | Control | 14596 | 7162 | 4790 | 5137 | 35.4% | 26.9% | 29.5% | 4.3% | 3.9% |
|  | IDA | Case | 503 | 262 | 0 | 503 | 22.5% | 43.3% | 26.4% | 4.8% | 3.0% |
|  |  | Control | 2998 | 1785 | 0 | 2998 | 42.0% | 23.5% | 26.1% | 5.2% | 5.2% |
|  | CKD-A | Case | 164 | 164 | 164 | 100 | 5.5% | 72.0% | 15.8% | 3.1% | 3.6% |
|  |  | Control | 2647 | 2647 | 2647 | 1184 | 24.3% | 33.8% | 32.9% | 3.4% | 5.6% |
|  | UA | Case | 914 | 575 | 245 | 536 | 21.1% | 60.1% | 12.3% | 3.4% | 3.1% |
|  |  | Control | 14596 | 7162 | 4790 | 5137 | 35.4% | 26.9% | 29.5% | 4.3% | 3.9% |
| Quantitative outcome approach | Hemoglobin | - | 28054 | 13554 | 7698 | 14232 | 40.2% | 24.6% | 27.1% | 4.1% | 4.1% |
|  | Serum VB12 | - | 27506 | 13328 | 7787 | 14032 | 40.2% | 24.5% | 27.3% | 4.0% | 4.0% |
|  | RBC folate | - | 29014 | 13977 | 8420 | 14769 | 39.7% | 24.9% | 27.2% | 4.1% | 4.1% |
|  | Serum Iron | - | 26564 | 14310 | 5850 | 13902 | 41.3% | 23.9% | 26.7% | 4.2% | 3.9% |

Note: The number of individuals was reported for each study with a breakdown by cohort, age, and ethnicity. QC: 948 quality control. IDA: iron deficiency anemia. CKD-A: chronic kidney disease-related anemia. UA: unexplained 949 anemia. EWAS: environment-wide association study. 950

### Survey Weights

Survey variable weights were incorporated in the EWAS regression models by CLARITE using the *survey* package in R to generate representative results for the US population^20^ (https://github.com/cran/survey). Two survey matrices were generated with and without replication by assigning appropriate weights to the variables. Variables in the discovery dataset were assigned the four-year weights. Variables in the replication cohort were assigned the two-year weights divided in half, as recommended by NHANES (Centers for Disease Control and Prevention). For studies without a sufficient sample size (< 200 people in either discovery or replication cohort), the discovery and replication cohorts were merged, and eight-year survey weights were used. This provided sufficient sample size and statistical power to identify the significant associations for these phenotypes. The corresponding eight-year weight was calculated and assigned to each variable based on the survey cycle: 1) half of the four-year weight for survey cycles one and two, and 2) a quarter of the two-year weight for survey cycles three and four (Centers for Disease Control and Prevention).

### Environment-wide Association Studies (EWAS)

All EWAS were performed using CLARITE and included the survey weights (Lucas et al. 2019). The case-control EWAS were performed using a binomial regression model, while a generalized linear model was used for the quantitative EWAS. All EWAS were adjusted for age, sex (male or female), race/ethnicity (African American, Caucasian American, Mexican American, other Hispanic, or other ethnicities), socioeconomic status (categorized as level 0, 1, or 2), and survey year (categorized as 1 for 1999−2000, 2 for 2001−2002, 3 for 2003−2004, or 4 for 2005−2006). Additional covariates were used to adjust for confounding variables for specific outcomes (Table 4). Cotinine levels were added as a covariate in all case-control EWAS and in the hemoglobin quantitative EWAS in adults because of the association between cotinine in serum and hemoglobin A_1c_ in the NHANES cohort (Clair et al. 2011). Iron supplement intake status was added as a covariate in the hemoglobin quantitative EWAS. Dietary vitamin B12 and vitamin B12 supplement intake were added as covariates in the vitamin B12 deficiency anemia quantitative EWAS. Folic acid supplement intake was added as a covariate in the folate deficiency anemia quantitative EWAS. Pregnancy status was added as a covariate in the IDA case-control EWAS.

For categorical variables, an additional likelihood ratio test (LRT) was performed to report an LRT p-value between the full model (model with the interest predictor and covariates) and the reduced model (model with the covariates and without the interest predictor). Significant exposures based on the discovery EWAS with a false discovery rate (FDR) < 10% were considered for replication, adjusting for the same covariates (Benjamini and Hochberg 1995). Exposures with a Bonferroni-corrected p < 0.05 in the replication cohort were considered replicated. Manhattan plots were created to display the EWAS results, ordered by variable category with labels for Bonferroni-corrected significant levels (alpha = 0.05), based on the number of discovery tests, and replicating significant associations labeled. A Hudson plot was generated using the *hudson* package (Lucas et al. 2022) in R to compare the results between EWAS for iron concentrations from the frozen samples and refrigerated samples in children to evaluate patterns across inconsistent measurements from two storage methods. The circular bar plots were plotted by using *ggplot2* package (Wickham 2009) and the chord diagrams were plotted by using *circlize* package (Gu et al. 2014) for summarizing the significant results in R. Cytoscape (version 3.8.2.) was used for the summary network plot (Shannon et al. 2003).

## Results

Across all the EWAS, we identified 265 significant results in the discovery cohort, and 106 were replicated (19 for children; 87 for adults). Top results are summarized in Figure 2A for children and Figure 2B for adults. Environmental exposures were classified by NHANES as nutrients, food component recall, supplements, pesticides, heavy metals, and smoking behaviors. In children, nutrients from blood tests or dietary recalls (11), supplement use (7), and heavy metals (1) were common exposures identified across anemia EWAS (Figure 2C). In particular, heavy metals, pesticides, dialkyl, and phthalate were also significantly associated with multiple anemias in children. In adults, key exposure categories across the anemia EWAS included nutrients (65), lifestyle factors (9, e.g., alcohol use or smoking), supplement use (7), heavy metals (3), phthalates (2), and birth control drug use (1) (Figure 2D).

**Figure 2.**
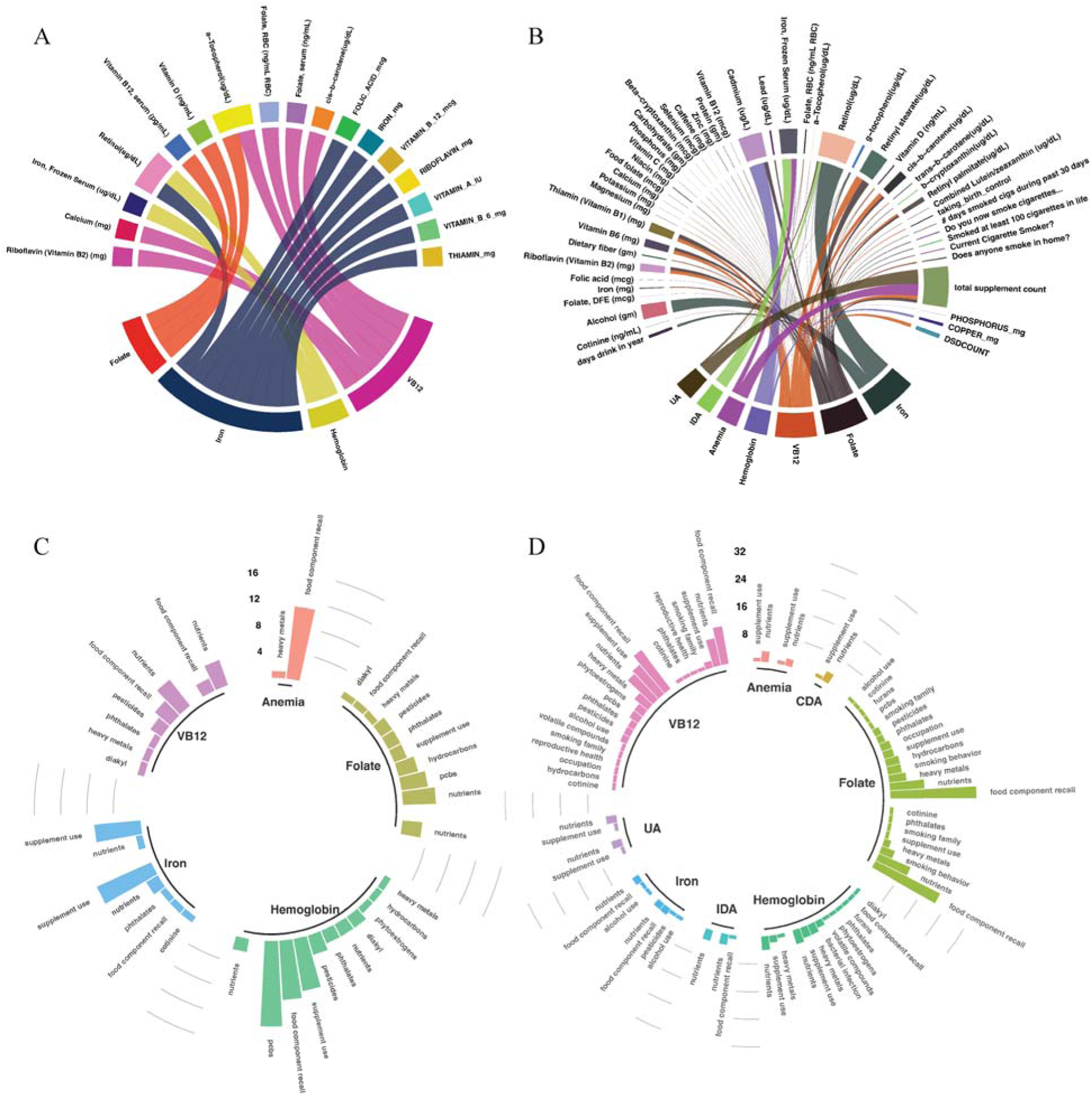
Replicating exposures associated with anemia in children and adults. The exposures compared between the anemic and control participants from discovery results with an FDR < 10% were considered for the replication with adjustment for the same covariates. The significant results from replications were based on a Bonferroni-adjusted p-value < 0.05. The chord diagrams show the replicating EWAS results for anemia, folate, hemoglobin, iron, and VB12 by exposures in children (A) and the replicating EWAS results for anemia, CDA, folate, hemoglobin, IDA, iron, UA, and VB12 by exposures in adults (B). The width of the link represents the effect size (β) of the association between exposure and phenotype. The circular bar plots show the replicating EWAS results for anemia, folate, hemoglobin, iron, and VB12 by exposure category in children (C) and the replicating EWAS results for anemia, CDA, folate, hemoglobin, IDA, iron, UA, and VB12 by exposure category in adults (D). Histogram graphs for each exposure category represent the numbers of identified significant exposures from discovery and replication dataset. Note: Folate, RBC (ng/mL RBC): folate in red blood cell in ng/mL. FOLIC_ACID_mcg: supplement obtained folic acid in µg. IRON_mg: supplement obtained iron in mg. VITAMIN_B_12_mcg: supplement obtained vitamin B12 in µg. RIBOFLAVIN_mg: supplement obtained riboflavin in mg. VIAMIN_A_IU: supplement obtained vitamin A in international units (IU). VITAMIN_B_6_mg: supplement obtained vitamin B6 in mg. THIAMIN_mg: supplement obtained thiamin in mg. VB12: vitamin B12. Folate, DFE: dietary folate equivalent intake in µg. Folate, RBC (ng/mL RBC): folate in red blood cell in ng/mL. PHOSPHORUS_mg: supplemental phosphorus in mg. COPPER_mg: supplemental copper in mg. DSDCOUNT: total number of dietary supplements taken. UA: unexplained anemia. IDA: iron deficiency anemia. VB12: Vitamin B12.

### Results of Case-Control EWAS

NHANES cohorts were divided into participants with anemia and healthy participants referencing their age, sex, and hemoglobin concentration, according to the WHO criteria (WHO 2011). The case-control EWAS were performed for general anemia, IDA, CKD-A, and UA in children (age < 15 years old) and adults (age ≥ 15 years). CKD-A analysis was restricted to adults because no children with CKD-A were identified in the NHANES dataset and chronic kidney disease in children is rarely reported in many countries (Becherucci et al. 2016). A discovery analysis was performed for a limited number of children with and without UA in a single case-control study. However, none of the variables was replicated under case-control EWAS for children.

Among adults, three exposures replicated for general anemia: frozen serum iron concentration (μg/dL; discovery: β = −0.053, FDR p: 8.108 × 10^−7^; replication: β = −0.079, Bonferroni p: 8.048 × 10^−9^), RBC folate (ng/mL; discovery: β = 0.005, FDR p: 6.59 × 10^−6^; replication: β = 0.004, Bonferroni p: 7.60 × 10^−7^), and total supplement count (total counts for all supplements that the individual used within 30 days; discovery: β = 0.276, FDR p: 5.18 × 10^−6^; replication: β = 0.205, Bonferroni p: 1.47 × 10^−2^) (Figure 3).

**Figure 3.**
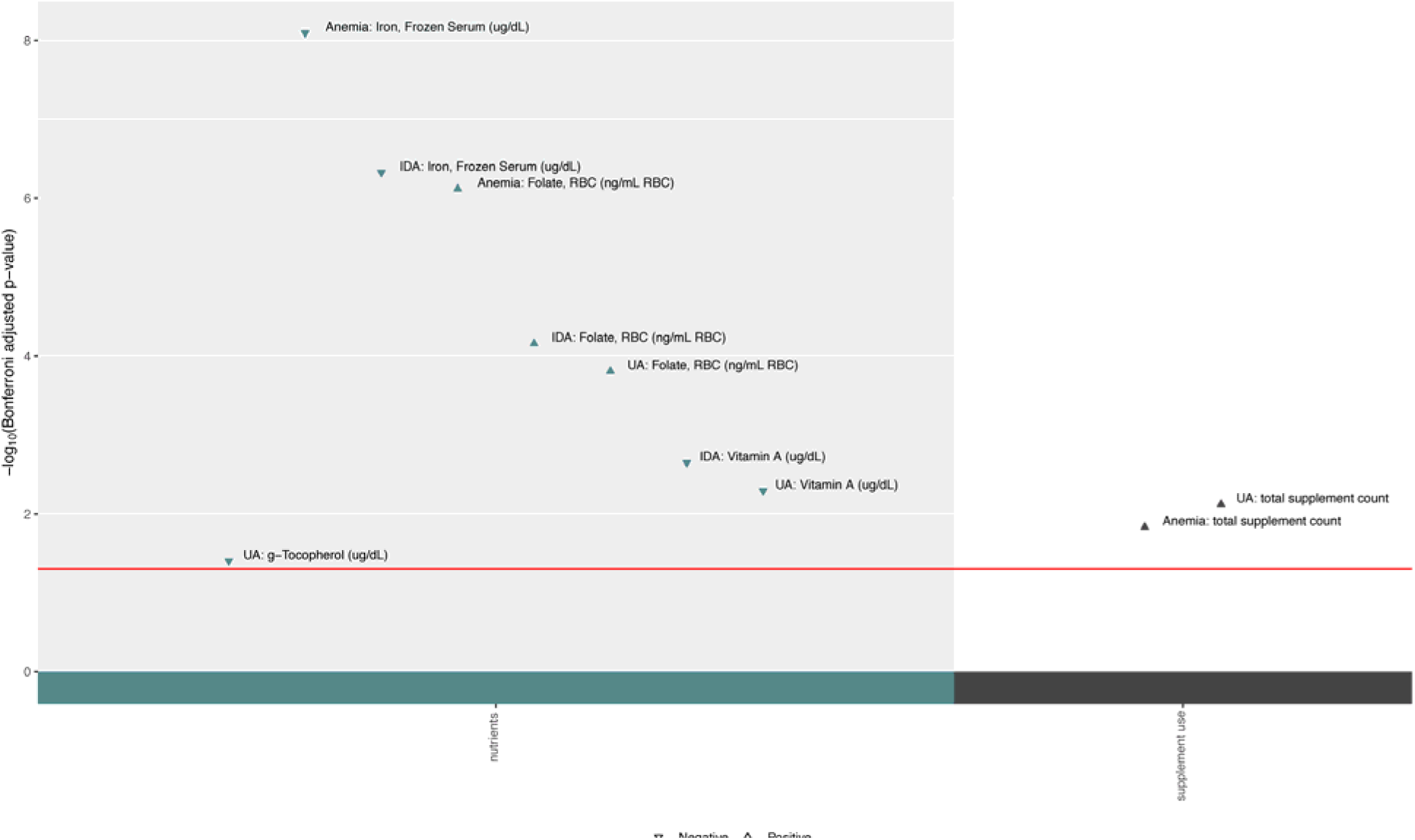
Manhattan plot for EWAS results from case-control in adults. The statistical comparison was made between the participants with anemia and the healthy participants by using binomial in GLM with adjustment for age, sex, race, socioeconomic status, survey year, and cotinine among young adults and adults (> 15 years old). The LRT was performed to compare the model with and without each categorical environmental factor by adjusting the same set of covariates. The x-axis shows the NHANES classified exposure categories, and the y-axis shows the -log_10_(Bonferroni adjusted p-value). The replicated results meeting discovery significance (FDR significant p < 0.1) and replication significance (Bonferroni significant p < 0.05) were represented by triangle. The direction of the effect size (β) was presented by the up-pointing triangle for positive effect and down-pointing triangle for negative effect. Note: Anemia: general anemia; IDA: iron deficiency anemia; UA: unexplained anemia; g−Tocopherol: gamma−Tocopherol; RBC: red blood cells.

The case-control EWAS for IDA was performed only for female adults since the males and children with IDA were not available in the study. Two of the four results significant in the discovery cohort were replicated in female adults: frozen serum iron concentration (μg/dL; discovery: β = −0.167, FDR p: 1.59 × 10^−5^; replication: β = −0.159, Bonferroni p: 4.73 × 10^−7^) and RBC folate (ng/mL RBC; discovery: β = 0.007, FDR p: 3.99 × 10^−4^; replication: β = 0.005, Bonferroni p: 6.95 × 10^−5^) (Figure 3).

For UA, all four variables meeting the discovery threshold were replicated, including RBC folate (ng/mL RBC; discovery: β = 0.003, FDR p: 1.30 × 10^−3^; replication: β = 0.003, Bonferroni p: 1.556 × 10^−4^), serum retinol (μg/dL; discovery: β = -0.018, FDR p: 0.039; replication: β = -0.025, Bonferroni p: 0.005), serum γ-tocopherol (μg/dL; discovery: β = -0.004, FDR p: 0.055; replication: β = -0.002, Bonferroni p: 0.039), and total supplement count (discovery: β = 0.255, FDR p: 1.30 × 10^−3^; replication: β = 0.230, Bonferroni p: 7.57 × 10^−3^) (Figure 3).

There were few CKD-A cases (272 cases; 98 for discovery in 1999−2002, and 174 for replication in 2003−2006; Table 2); therefore, a case-control analysis for all adults in a single discovery group without replication was performed. Three exposures were significant, including serum retinol (μg/dL; discovery: β = 0.059, Bonferroni p: 4.10 × 10^−21^), RBC folate (ng/mL RBC; discovery: β = 0.058, Bonferroni p: 1.31 × 10^−14^), and total supplement count (discovery: β = 0.215, Bonferroni p: 7.47 × 10^−5^) (Figure S1).

### EWAS results for hemoglobin concentration

Hemoglobin concentration levels were used to evaluate significant exposure associations. Forty-two exposures in the discovery were considered for replication in children. Two replicated: frozen serum iron concentration (μg/dL; discovery: β = 0.005, FDR p: 7.20 × 10^−5^; replication: β = 0.006, Bonferroni p: 1.91 × 10^−5^) and serum retinol (μg/dL; discovery: β = 0.028, FDR p: 3.01 × 10^−6^; replication: β = 0.025, Bonferroni p: 2.58 × 10^−3^) (Figure 4).

**Figure 4.**
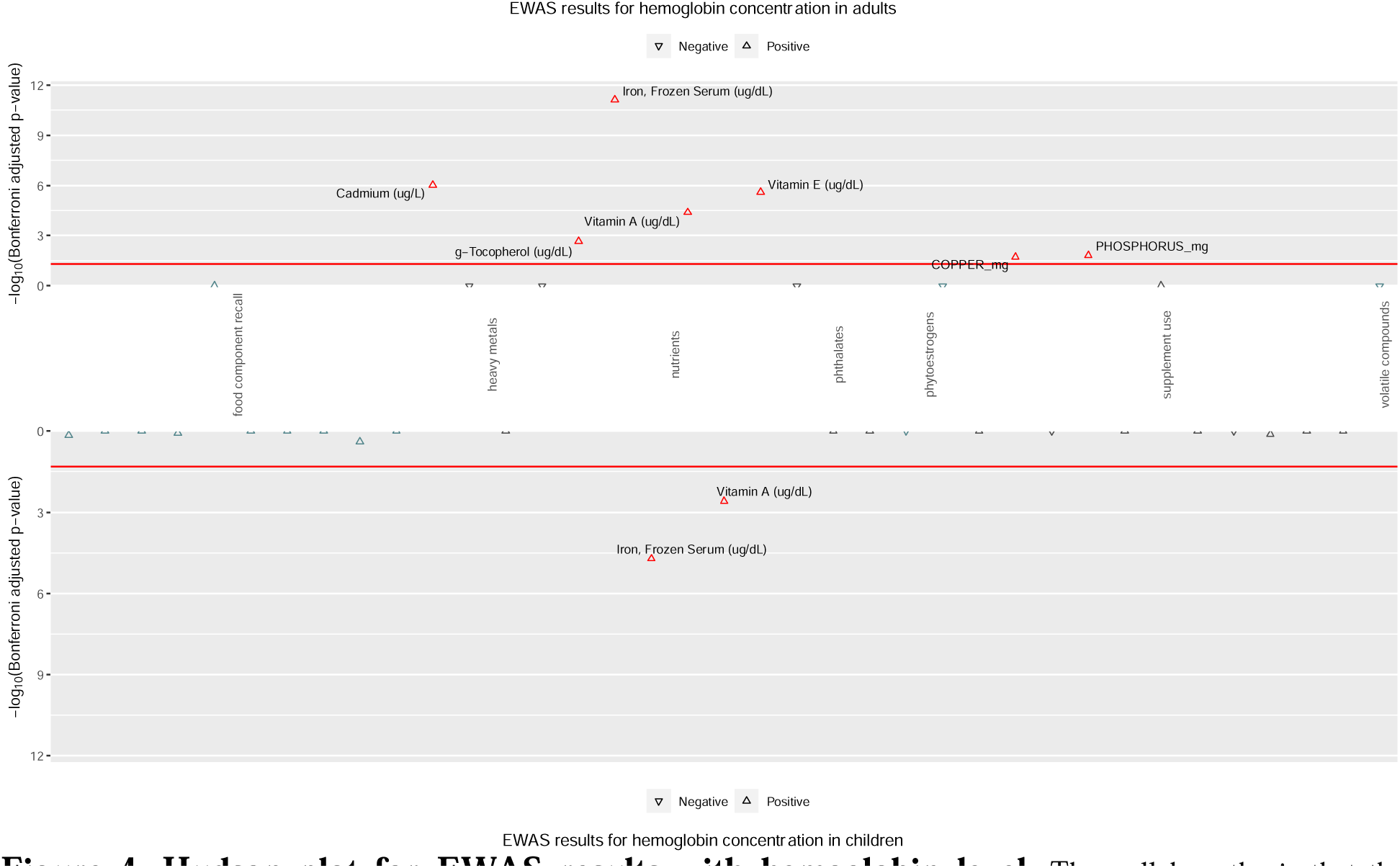
Hudson plot for EWAS results with hemoglobin level. The null hypothesis that the coefficient is equal to zero (no effect) was tested in a generalized linear model by using the hemoglobin level as the outcome and adjusting for age, sex, race, socioeconomic status, survey year, and iron supplement intakes among adults (top panel) and children (bottom panel). The LRT was performed to compare the model with and without each categorical environmental factor by adjusting the same set of covariates. The x-axis shows the NHANES classified exposure categories, and the y-axis shows the -log_10_(Bonferroni adjusted p-value). The significant replicated results meeting discovery significance (FDR significant p < 0.1) and replication significance (Bonferroni significant p < 0.05; red line) were highlighted by red triangle. The direction of the effect size (β) was presented by the up-pointing triangle for positive effect and down-pointing triangle for negative effect. Note: g−Tocopherol: γ-Tocopherol.

In adults, seven exposures were replicated out of 18 exposures meeting the discovery threshold. Frozen serum iron concentration (μg/dL; discovery: β = 0.008, FDR p: 3.36 × 10^−10^; replication: β = 0.011, Bonferroni p: 7.47 × 10^−12^), blood cadmium (μg/L; discovery: β = 0.294, FDR p: 8.56 × 10^−5^; replication: β = 0.258, Bonferroni p: 9.55 × 10^−7^), serum α-tocopherol (μg/L; discovery: β = 0.0002, FDR p: 1.91 × 10^−3^; replication: β = 0.0002, Bonferroni p: 2.57 × 10^−6^), serum retinol (μg/dL; discovery: β = 0.009, FDR p: 6.29 × 10^−6^; replication: β = 0.009, Bonferroni p: 4.05 × 10^−5^), serum γ-tocopherol (μg/dL; discovery: β = 4.57 × 10^−4^, FDR p: 0.068; replication: β = 6.11 × 10^−4^, Bonferroni p: 4.26 × 10^−4^), phosphorus (mg; discovery: β = 4.15 × 10^−4^, FDR p: 0.068; replication: β = 5.49 × 10^−5^, Bonferroni p: 0.015), and copper (mg; discovery: β = 0.074, FDR p: 0.019; replication: β = 0.04, Bonferroni p: 0.019) were significantly associated with hemoglobin concentration (Figure 4).

### EWAS results for serum vitamin B12 concentration

Serum Vitamin B12 levels were used to classify vitamin B12 deficiency anemia. Since there were few cases of vitamin B12 deficiency anemia, vitamin B12 serum levels were used as a quantitative outcome to assess significant exposures. In children, six of 13 environmental exposures meeting the discovery threshold were replicated, including RBC folate (ng/mL RBC; discovery: β = 0.0008, FDR p: 3.41 × 10^−4^; replication: β = 0.0008, Bonferroni p: 1.90 × 10^−5^), serum folate (ng/ml; discovery: β = 0.012, FDR p: 5.23 × 10^−8^; replication: β = 0.006, Bonferroni p: 8.45 × 10^−5^), dietary riboflavin (mg; discovery: β = 0.039, FDR p: 0.006; replication: β = 0.073, Bonferroni p: 6.05 × 10^−4^), dietary calcium (mg; discovery: β = 6.25 × 10^−5^, FDR p: 0.024; replication: β = 1.29 × 10^−4^, Bonferroni p: 7.73 × 10^−4^), serum α-tocopherol (μg/L; discovery: β = 0.0002, FDR p: 1.53 × 10^−4^; replication: β = 0.0003, Bonferroni p: 1.66 × 10^−3^), and cis-β-carotene (μg/dL; discovery: β = 0.108, FDR p: 0.097; replication: β = 0.066, Bonferroni p: 5.21 × 10^−3^) (Figure 5).

**Figure 5.**
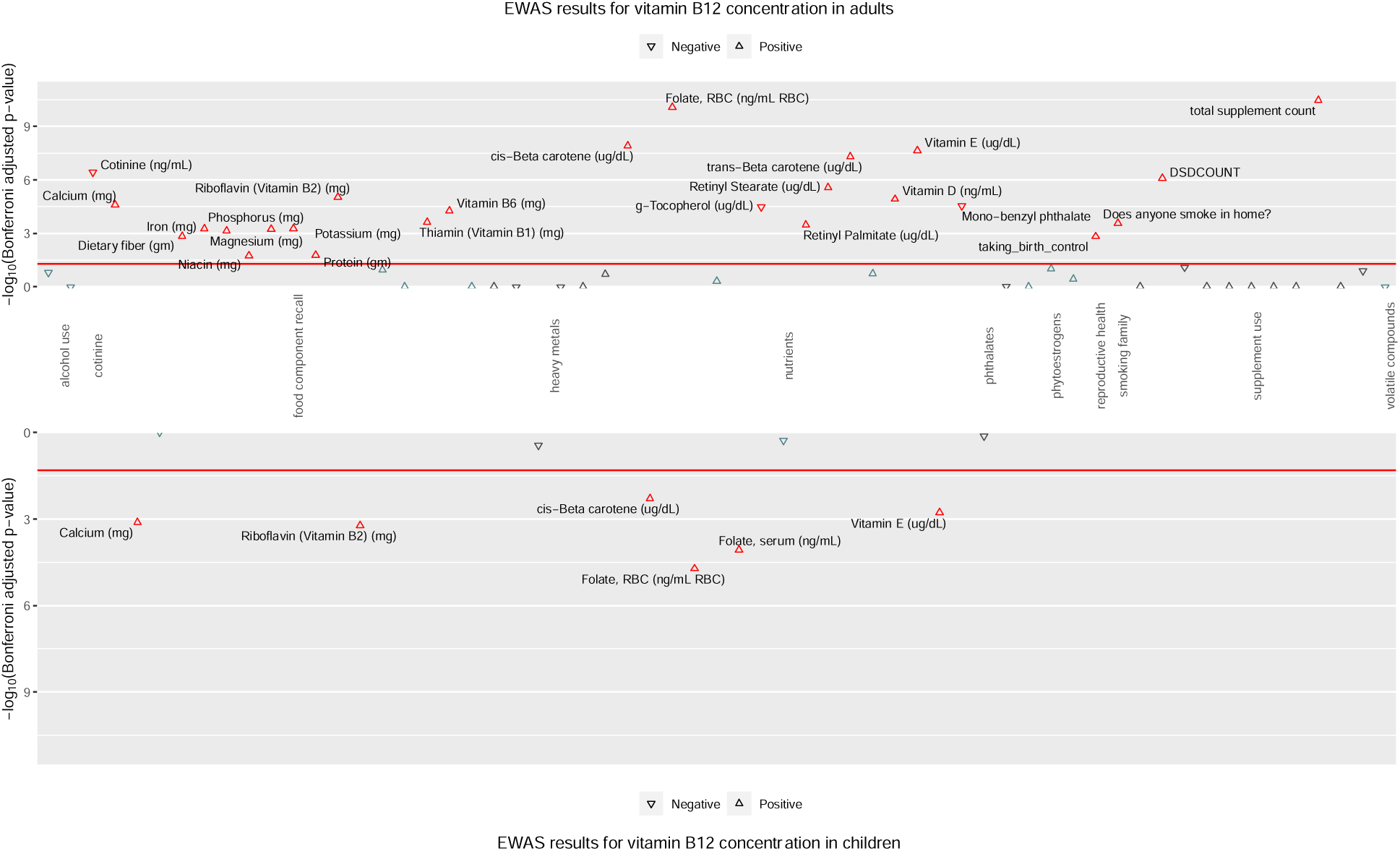
Hudson plot for EWAS results with serum vitamin B12 level. The null hypothesis that the coefficient is equal to zero (no effect) was tested in a generalized linear model by using the serum vitamin B12 level as the outcome and adjusting for age, sex, race, socioeconomic status, survey year, dietary intake of vitamin B12, and vitamin B12 supplement intakes among adults (top panel) and children (bottom panel). The LRT was performed to compare the model with and without each categorical environmental factor by adjusting the same set of covariates. The x-axis shows the NHANES classified exposure categories, and the y-axis shows the - log_10_(Bonferroni adjusted p-value). The significant replicated results meeting discovery significance (FDR significant p < 0.1) and replication significance (Bonferroni significant p < 0.05; red line) were highlighted by red triangle. The direction of the effect size (β) was presented by the up-pointing triangle for positive effect and down-pointing triangle for negative effect. Note: g−Tocopherol: γ-Tocopherol; RBC: red blood cell; DSDCOUNT: the counts of supplements used in 30 days.

Twenty-five dietary variables of 59 exposures significant in the discovery cohort were replicated for serum vitamin B12 concentration levels in adults. The top results included total supplement count (discovery: β = 0.050, FDR p: 1.33 × 10^−8^; replication: β = 0.00007, Bonferroni p: 3.38 × 10^−11^), RBC folate (ng/mL RBC; discovery: β = 0.0008, FDR p: 2.72 × 10^−7^; replication: β = 0.001, Bonferroni p: 8.49 × 10^−11^), cis-β-carotene (μg/dL; discovery: β = 0.090, FDR p: 0.033; replication: β = 0.071, Bonferroni p: 1.25 × 10^−8^), serum α-tocopherol (μg/dL; discovery: β = 1.34 × 10^−4^, FDR p: 9.21 × 10^−7^; replication: β = 1.75 × 10^−4^, Bonferroni p: 2.29 × 10^−8^), cis-β-carotene (μg/dL; discovery: β = 0.005, FDR p: 0.044; replication: β = 0.005, Bonferroni p: 5.06 × 10^−8^), and serum cotinine (ng/mL; discovery: β = -2.05 × 10^−4^, FDR p: 0.006; replication: β = -3.13 × 10^−4^, Bonferroni p: 5.06 × 10^−8^) (Figure 5). Other replicating exposures include: Dietary intake of riboflavin, calcium, magnesium, iron, fiber, thiamine, potassium, protein, phosphorus, vitamin B6, niacin, blood level of retinyl palmitate, retinyl Stearate, vitamin D, γ-tocopherol, urine measurement of mono-benzyl phthalate, and survey questions on whether they were taking birth control drugs, whether any family member were smoking in home, and total number of dietary supplements taken were also significantly associated with serum vitamin B12 level.

### EWAS results for folate levels in RBC

Since there were few cases of folate deficiency anemia among children and adults (16 cases from 1999–2006; Table 2), we performed an EWAS with RBC folate as a quantitative measure. Three of 21 variables considered replicated in children, which are serum vitamin B12 (pg/mL; discovery: β = 0.0002, FDR p: 9.30 × 10^−5^; replication: β = 0.0002, Bonferroni p: 2.66 × 10^−5^), serum vitamin D (ng/mL; discovery: β = 0.005, FDR p: 0.097; replication: β = 0.004, Bonferroni p: 3.76 × 10^−3^), and serum α-tocopherol (μg/dL; discovery: β = 0.0001, FDR p: 0.070; replication: β = 0.0001, Bonferroni p: 0.035) (Figure 6).

**Figure 6.**
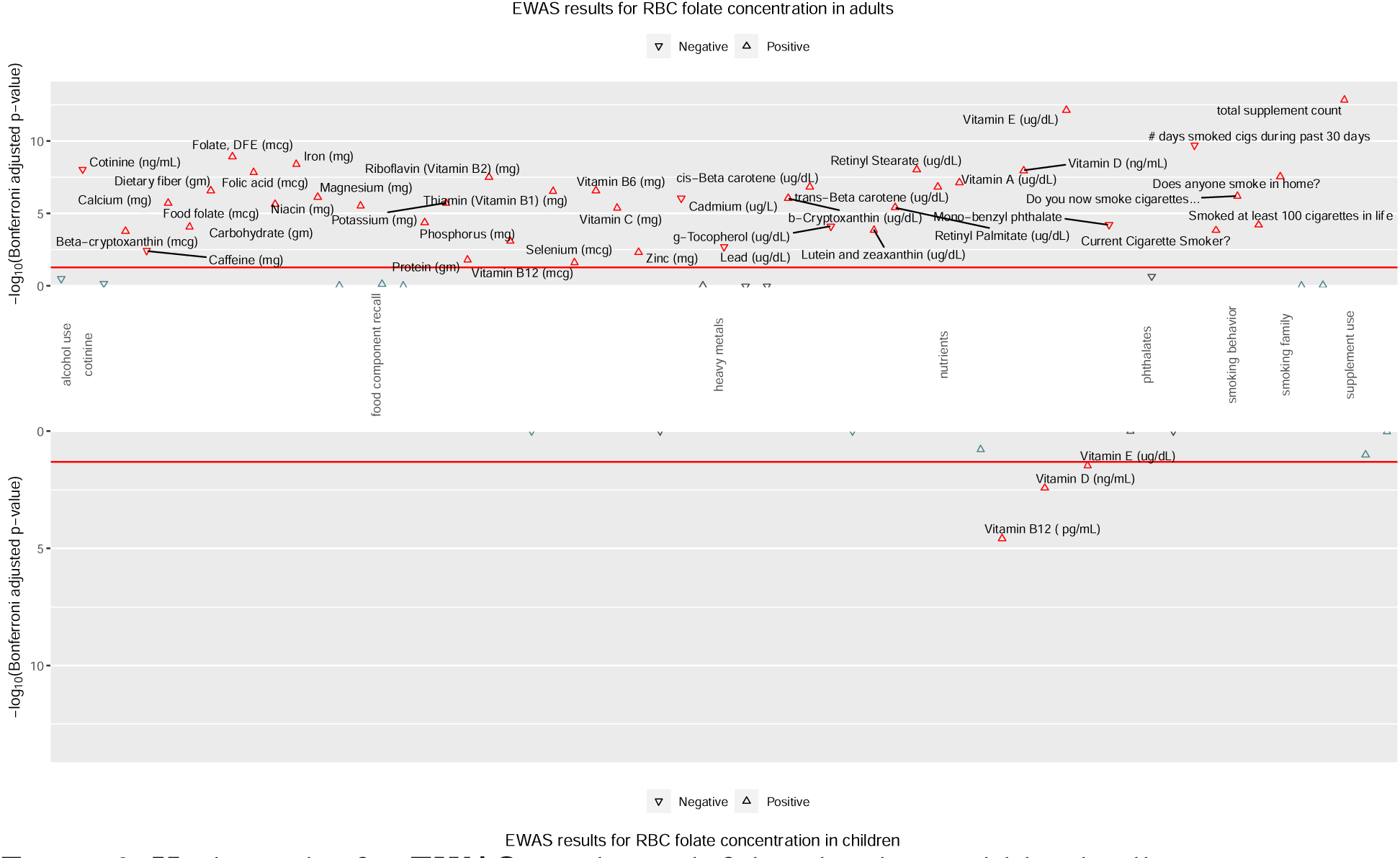
Hudson plot for EWAS results with folate level in red blood cell. The null hypothesis that the coefficient is equal to zero (no effect) was tested in a generalized linear model by using the folate level in red blood cell as the outcome and adjusting for age, sex, race, socioeconomic status, survey year, and folic acid supplement intakes among adults (top panel) and children (bottom panel). The LRT was performed to compare the model with and without each categorical environmental factor by adjusting the same set of covariates. The x-axis shows the NHANES classified exposure categories, and the y-axis shows the -log_10_(Bonferroni adjusted p-value). The significant replicated results meeting discovery significance (FDR significant p < 0.1) and replication significance (Bonferroni significant p < 0.05; red line) were highlighted by red triangle. The direction of the effect size (β) was presented by the up-pointing triangle for positive effect and down-pointing triangle for negative effect. Note: g−Tocopherol: γ-Tocopherol; b-Cryptoxanthin: β-Cryptoxanthin; mcg: μg; DFE: dietary folate equivalent.

In adults, 41 exposures of the 64 considered replicated. Multiple exposures were related to smoking status and dietary measures including metal ions, vitamin Bs, and fiber with several serum measurements. The top exposures included total supplement count (discovery: β = 0.029, FDR p: 2.43 × 10^−4^; replication: β = 0.053, Bonferroni p: 1.43 × 10^−13^), serum α-tocopherol (μg/L; discovery: β = 0.0001, FDR p: 4.34 × 10^−6^; replication: β = 0.0002, Bonferroni p: 7.47 × 10^−13^), number of days smoked cigarette during past 30 days (discovery: β = -0.004, FDR p: 0.012; replication: β = -0.005, Bonferroni p: 1.95 × 10^−10^), dietary folate equivalents (folate, DFE, mg; discovery: β = 0.0001, FDR p: 0.008; replication: β = 0.0002, Bonferroni p: 1.21 × 10^−9^), dietary intake of iron (mg; discovery: β = 0.005, FDR p: 4.41 × 10^−8^; replication: β = 0.009, Bonferroni p: 4.01 × 10^−9^), and serum cotinine (ng/mL; discovery: β = −0.0005, FDR p: 9.22 × 10^−7^; replication: β = −0.0005, Bonferroni p: 8.97 × 10^−9^) (Figure 6). The remaining replicated exposures included nutrients from dietary intake recall survey including vitamin B1, riboflavin, vitamin B6, vitamin B12, vitamin C, magnesium, niacin, calcium, potassium, phosphorus, zinc, selenium, carbohydrate, food folate, folic acid, fiber, protein, caffeine, β-cryptoxanthin and from serum measurements for β-cryptoxanthin, γ-tocopherol, vitamin D, retinyl palmitate, retinyl stearate, cis-β-carotene, trans-β-carotene, lutein and zeaxanthin, and retinol. Additional replicated exposures included heavy metals, cadmium, lead, mono-benzyl phthalate, and whether an individual smoked at least 100 cigarettes in their lifetime, currently smokes, and smokes at home.

**Figure 6.**
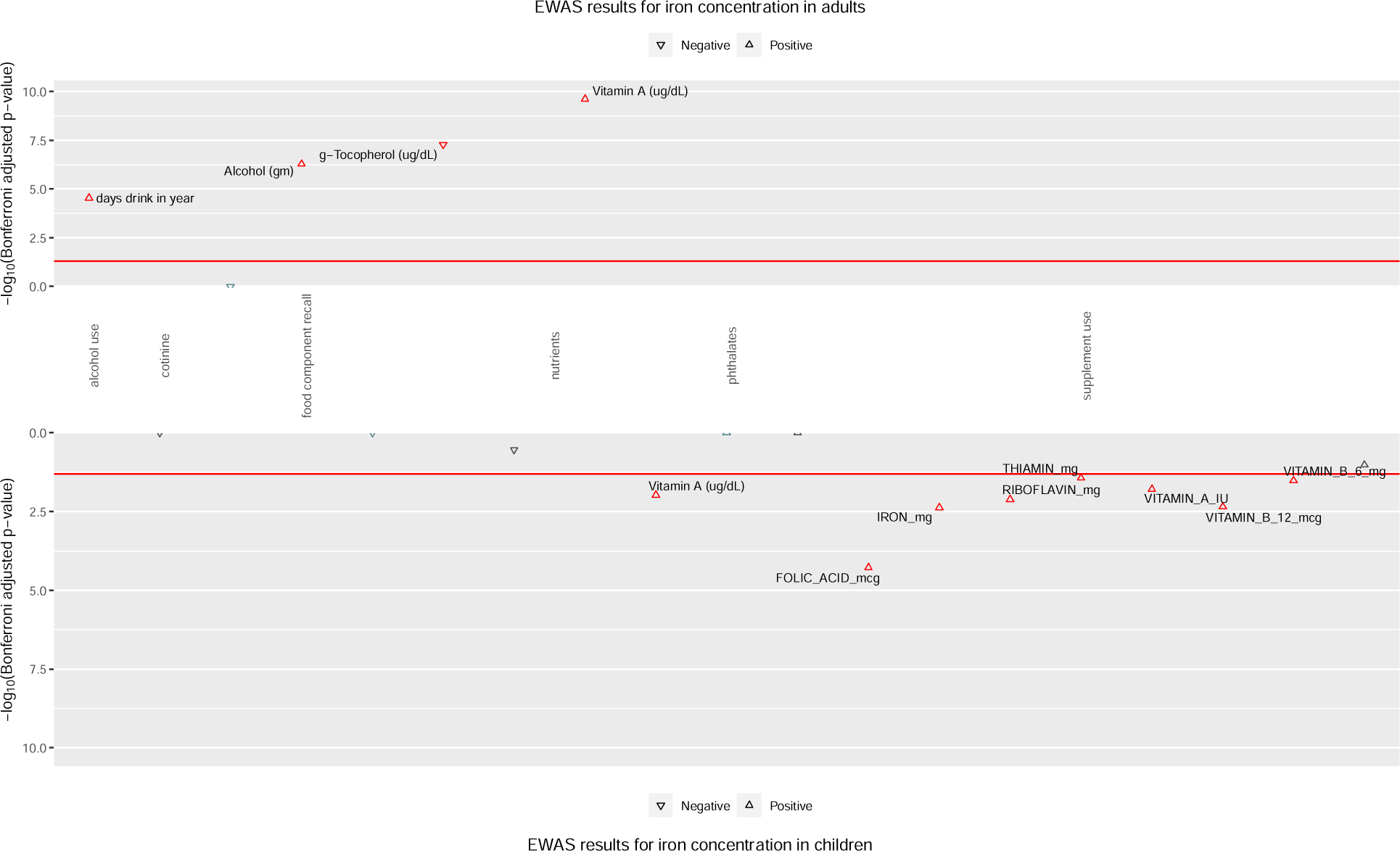
Hudson plot for EWAS results with serum iron level. The null hypothesis that the coefficient is equal to zero (no effect) was tested in a generalized linear model by using the serum iron concentration as the outcome and adjusting for age, sex, race, socioeconomic status, and survey year among adults (top panel) and children (bottom panel). The LRT was performed to compare the model with and without each categorical environmental factor by adjusting the same set of covariates. The x-axis shows the NHANES classified exposure categories, and the y-axis shows the -log_10_(Bonferroni adjusted p-value). The significant replicated results meeting discovery significance (FDR significant p < 0.1) and replication significance (Bonferroni significant p < 0.05; red line) were highlighted by red triangle. The direction of the effect size (β) was presented by the up-pointing triangle for positive effect and down-pointing triangle for negative effect. Note: g−Tocopherol: γ-Tocopherol.

### EWAS results for serum iron concentration

In the NHANES dataset, serum iron concentrations were measured from their frozen blood samples and/or refrigerated blood samples. Some individuals had only a single measurement for the serum iron concentration from either the frozen or refrigerated blood samples. Hence, we performed the association studies by 1) using the frozen serum iron samples and 2) using the combined values for serum iron from frozen and refrigerated samples for children since the measurements were inconsistent. Either approach gave similar results (Figure S2); thus, we report the significant results from frozen serum iron as the quantitative outcome. In the child cohort, eight exposures of 14 considered replicated including supplements of folic acid (μg; discovery: β = 0.00008, FDR p: 1.83 × 10^−9^; replication: β = 0.006, Bonferroni p: 5.36 × 10^−5^), iron (mg; discovery: β = 0.002, FDR p: 1.83 × 10^−9^; replication: β = 0.132, Bonferroni p: 4.24 × 10^−3^), vitamin A (international unit (IU); discovery: β = 0.000006, FDR p: 1.12 × 10^−10^; replication: β = 0.0005, Bonferroni p: 1.63 × 10^−2^), vitamin B6 (mg; discovery: β = 0.016, FDR p: 0.005; replication: β = 0.276, Bonferroni p: 3.01 × 10^−2^), vitamin B12 (μg; discovery: β = 0.005, FDR p: 5.97 × 10^−7^; replication: β = 0.253, Bonferroni p: 4.46 × 10^−3^), riboflavin (mg; discovery: β = 0.018, FDR p: 0.0002; replication:, β = 0.388, Bonferroni p: 7.57 × 10^−3^), thiamine (mg; discovery: β = 0.022, FDR p: 0.009; replication:, β = 0.363, Bonferroni p: 0.037) and serum vitamin A level (μg/dL; discovery: β = 1.000, FDR p: 3.56 × 10^−6^; replication: β = 0.849, Bonferroni p: 1.04 × 10^−2^) (Figure 7).

In adults, we found that four associations out of six variable meeting discovery threshold with serum iron concentration including serum vitamin A (μg/dL; discovery: β = 0.302, FDR p: 8.24 × 10^−8^; replication: β = 0.370, Bonferroni p: 2.40 × 10^−10^), serum γ-tocopherol (μg/dL; discovery: β = −0.028, FDR p: 9.10 × 10^−7^; replication: β = −0.027, Bonferroni p: 5.17 × 10^−8^), alcohol consumption (g; discovery: β = 0.127, FDR p: 2.74 × 10^−6^; replication: β = 0.202, Bonferroni p: 5.17 × 10^−7^), and total drinking days per year (discovery: β = 0.039, FDR p: 0.007; replication: β = 0.041, Bonferroni p: 2.94 × 10^−5^) (Figure 7).

### The Interrelationships Between Anemia-Related Risk Factors

Most environmental risk factors for anemia shared by children and adults were nutrients determined by blood tests (9), nutrients from food component recall (2), and supplements (8) (Figure 8). For example, serum retinol concentration was significantly associated with serum iron concentration in children and adults, and both were significantly associated with the level of hemoglobin in both groups. Serum folate concentration, serum α-tocopherol (vitamin E), cis-β-carotene, use of calcium supplements, and use of supplemental riboflavin (vitamin B2) were significantly associated with serum vitamin B12 level. Also, vitamin D and serum α-tocopherol (vitamin E) were significantly associated with serum folate levels for both groups.

**Figure 8.**
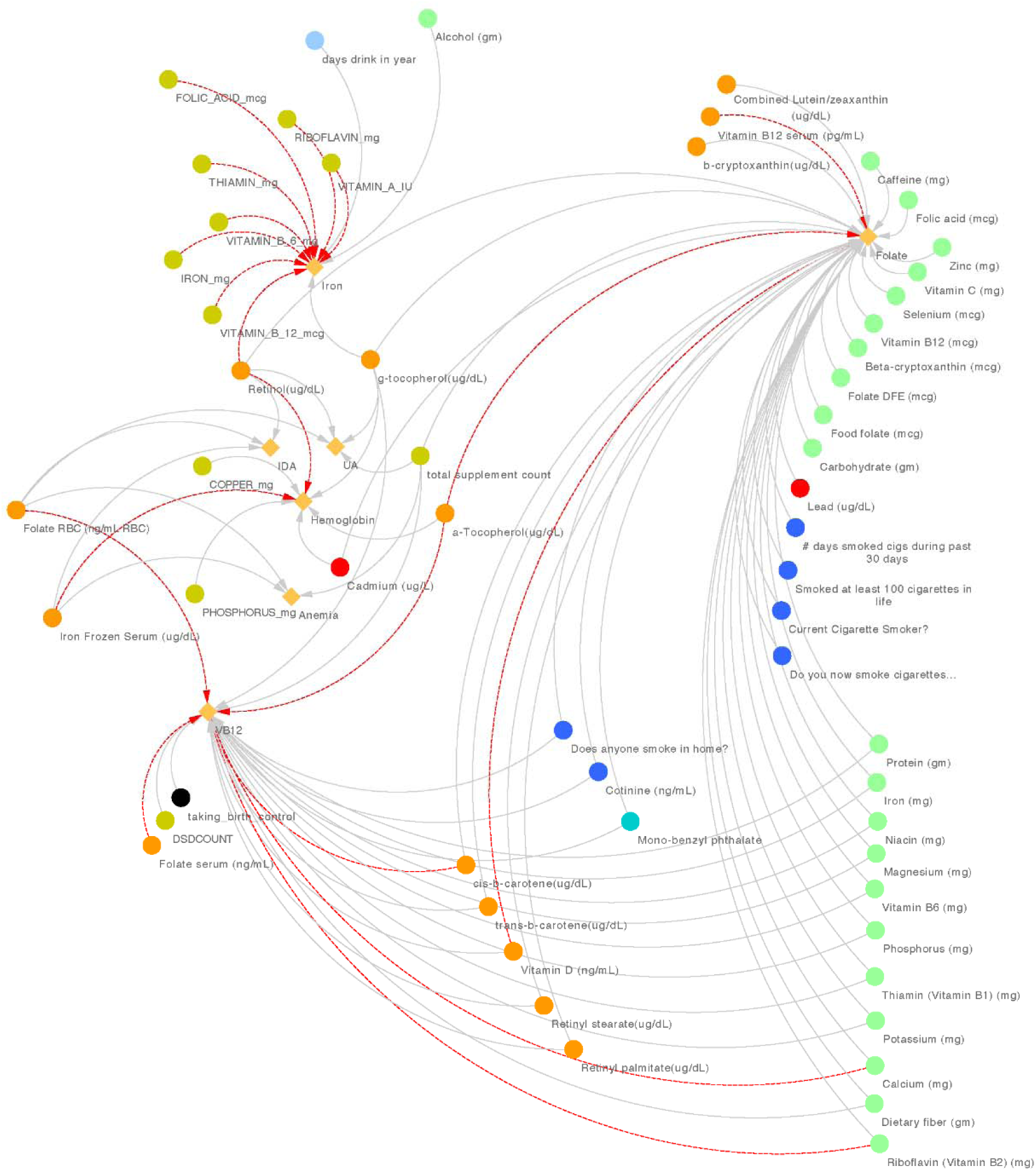
Network of the discoveries from the EWAS. The significant interactions with Bonferroni adjustment between phenotypes (a diamond) and environmental factors (a circle) are shown on the plot. The associations that were discovered are shown with a red dashed line for children and with a solid line for adults. The environmental factors are colored by their categories, including alcohol use (light blue), cotinine and smoking behaviors (dark blue), food component recall (light green), heavy metals (red), nutrients (orange), phthalates (cyan), and supplement use (dark green). Notes: RBC: red blood cell; DFE: dietary folate equivalent; DSDCOUNT: the counts of supplements used in 30 days.

Other than shared variables, the smoking behaviors and nutrients from blood tests and food component recall were the major categories of environmental risk factors associated with serum vitamin B12 concentration and serum folate concentration for adults. However, for children, only nutrients from blood tests were the major significant associations for these outcomes. The anemia phenotype that showed the greatest difference in environmental exposures between children and adults was serum iron levels. While most significant environmental exposures associated with serum iron concentration in children fell in the supplement use category, alcohol consumption was the category with the most significant exposures in adults.

Additionally, 26 environmental factors were shared by different anemia phenotypes. For instance, serum α-tocopherol (vitamin E) was significantly associated with serum levels of vitamin B12 and folate for both children and adults. Serum retinol concentration was also significantly associated with serum iron concentrations and hemoglobin levels for children and adults. In general, nutrients, food component recall, smoking behaviors, and supplement use were commonly shared between serum levels of vitamin B12 and folate. Retinol, γ-tocopherol (vitamin E), α-tocopherol (vitamin E), folate from RBC, serum iron concentration, and total supplement counts were commonly associated with multiple anemias.

## Discussion

Here, we identified environmental risk factors for general anemia, IDA, vitamin B12 deficiency anemia, folate deficiency anemia, CDA, CKD-A, and UA through a series of EWAS in children and adults. The quantitative EWAS give more significant replicated results than the case-control EWAS for the same phenotype. The major exposures that were associated with different anemia phenotypes and were replicated were nutrients, heavy metals, and supplement use in both children and adults cohorts, and smoking behavior and alcohol consumption in adults only (Figure 21) (EPA 2017).

Vitamin A is essential for erythropoiesis through retinoid signaling and promotes iron absorption from food (Garcıia-Casal et al. 1998). Vitamin A deficiency decreases RBC production as well as iron absorption and leads to the development of anemias. Similarly, vitamin E deficiency leads to anemia, especially sickle cell anemia (Stone et al. 1990) and hemolytic anemia in premature infants (Lo et al. 1973; Oski and Barness 1967). We found that serum retinol (deficiency if < 0.35 μmol/L for children and if < 0.70 μmol/L for women; adequate: 0.1 to 1.0 μmol/g liver) (Tanumihardjo 2012) and serum α-tocopherol and γ-tocopherol (reference range: 0−17 years: 3.8−18.4 mg/L, ≥ 18 years: 5.5−17.0 mg/L) (Mayo Clinic Medical Laboratories) were associated with the majority of anemia phenotypes (Figures 4 and 5). Specifically, among children, vitamin A was significantly positively associated with level of iron, and the level of hemoglobin and α-tocopherol was positively associated with the level of vitamin B12 in serum and the level of folate in RBC. For adults, vitamin A was also significantly associated with most of the nutritional deficiency anemias, including IDA and folate deficiency anemia, and most of the non-nutritional deficiency anemias, including UA, level of hemoglobin, and level of serum iron. Moreover, the levels of vitamin B12 in serum and vitamin E, including α-tocopherol and γ-tocopherol, have significant associations with all anemia phenotypes except general anemia and IDA. These results are relevant to the roles of these vitamins in RBC production.

Cadmium is a heavy metal obtained from nickel-cadmium batteries, contaminated food, jewelry, tobacco, and pigments (WHO | Cadmium 2020). Serum cadmium (RfD in drinking water: 0.0005 milligrams per kilogram per day (mg/kg/day) and RfD for dietary exposure: 0.001 mg/kg/day) (USEPA 2016) was significantly associated with general anemia in pediatric subjects and low level of serum folate in adults in our study. Increased serum cadmium levels lead to depletion of aged erythrocytes and downregulation of erythropoietic activity in mice (Chatterjee and Saxena 2015). Hemolysis, iron accumulation, and lack of RBC production in rats are associated with hepatic, renal, and osteoid damage after cadmium exposure (Horiguchi et al. 2011). In Japan, chronic cadmium poisoning causes Itai-itai disease with renal anemia that is associated with downregulation of serum erythropoietin (Horiguchi et al. 1994). Patients with thalassemia major also accumulate cadmium (Bayhan et al. 2017). Thus, high cadmium interferes with erythrocyte production and iron metabolism and leads to anemia. However, we also found that blood cadmium and urinary cadmium levels were significantly associated with high levels of hemoglobin in adults. The level of cadmium in urine reflects the long-term exposure which will not suddenly increase after an acute exposure (Cadmium Toxicity: Clinical Assessment - Laboratory Tests | Environmental Medicine | ATSDR). One possible explanation is that the accumulation of urinary cadmium could be obtained from long-term tobacco smoke which would lead to an increase of hemoglobin (Malenica et al. 2017; Pedersen et al. 2019).

IDA was associated with serum RBC folate (deficiency if < 100 ng/mL) (World Health Organization 2015), and serum vitamin A levels (deficiency if < 0.35 μmol/L for children and if < 0.70 μmol/L for women; adequate: 0.1 to 1.0 μmol/g liver) (Tanumihardjo 2012). In clinical trials with adults, including pregnant women, an iron supplement with folate or vitamin A prevented IDA more effectively than iron supplementation alone (Juarez-Vazquez et al. 2002; Michelazzo et al. 2013). In a Chinese cohort, serum vitamin A was positively associated with hematocrit, another measurement of anemia, in an EWAS for cardiovascular disease (Zhong et al. 2016). In two randomized controlled trials, vitamin A supplements improved anemia and growth in children in Tanzania (Mwanri et al. 2000), and especially so for children with IDA in Malaysia (Al-Mekhlafi et al. 2013). These clinical studies are supported by data in rats demonstrating a role for serum vitamin A in the regulation of iron release from the liver (Staab et al. 1984).

Vitamin B12, vitamin B6, and riboflavin (vitamin B2) are required in folate metabolism to synthesize nucleic acids (Shane 2008) and promote methylation (Shane 2008) and erythropoiesis (Koury and Ponka 2004). In our study, vitamin B12 deficiency anemia and folate deficiency anemia were positively associated with other B vitamins and dietary minerals. Thus, a multi-vitamin B supplement could help prevent these two vitamin B-related anemias. Multi-vitamins increase hemoglobin in anemic children (Sivakumar et al. 2006) and hemoglobin and ferritin in pregnant women (Makola et al. 2003) reducing the risk of iron deficiency and anemia. Supplemental vitamin B6 is reported to reverse anemia in pregnant women (Hisano et al. 2010) and in a patient with microcytic sideroblastic anemia (Allain et al. 2019). The association between low level of vitamin B2 and anemia was also observed from a cross-sectional population study in a Chinese population with a five-year follow-up (Shi et al. 2014). Adequate vitamin B2 supplementation increased iron absorption from food (Mahabadi et al. 2020). However, exactly how these vitamins affect anemia remains unclear.

We identified lifestyle factors such as the use of cigarettes and alcohol as risk factors affecting RBC folate and serum iron levels in adults. Cotinine, the main component from cigarettes and metabolic product from nicotine, in serum and many smoking behaviors were inversely associated with RBC folate and serum vitamin B12 concentrations, and cigarette smoking has been shown to decrease the level of folate in RBCs (Nedrebø et al. 2003; Piyathilake et al. 1994).

We also found a positive association between alcohol consumption, including the amount consumed and the number of days of consumption/year, and serum iron concentration. Alcohol consumption increases serum iron levels and decreases hepatic iron levels and the production of ferritin by reducing the expression of hepcidin in the liver and serum (Varghese et al. 2016). Hepcidin is the key peptide hormone for regulating the absorption, storage, transport, and recycling of iron in mammals (Ganz 2016).

To prevent and treat common types of anemia, foods rich in iron, vitamin Bs, vitamin C, and corresponded supplements may help. For example, the NIH establishes that the best resources for obtaining iron are meat (red meat, poultry, and seafood), beans and bean products, dark green leafy vegetables, dried fruits, and iron- and folate-fortified products (Avoiding Anemia | NIH News in Health). For essential vitamins, vitamin A and vitamin C can be obtained from fruits, vegetables, milk and milk products; vitamin B6 and B12 can be obtained from protein food, milk, and milk products; vitamin E can be obtained from vegetables and oils; and folate and riboflavin can be obtained from grains and milk and milk products (Whitney and Rolfes 2016). On the other hand, solid fats and/or added sugar must be limited while people consume food enriched with the recommended essential nutrients. Other than food enriched with vitamins, healthy lifestyles, including quitting or reducing the use of tobacco and alcohol, prevents the development of anemias.

There are some limitations to this study. First, the statistical power for the case-control studies for CDA, CKD-A, vitamin B12 deficiency anemia, and folate deficiency anemia was weak because the sample sizes were small. Quantitative variables for these phenotypes may not reflect the differences between the cases and controls. The classification of anemia from chronic disease/inflammation lacks source specificity, which could be supplied by a clinical diagnosis. Although medical records were available, only general chronic disease and chronic kidney inflammation were classifiable from NHANES data using the GFR and TBI with serum iron levels (Astor et al. 2002; Coresh et al. 2007; Gupta et al. 2017; Guralnik et al. 2004; Park and Eicher-Miller 2014; Patel et al. 2009; Weiss et al. 2019a). If they were available, organ-specific inflammation measurements could identify anemias originating from immune cells, liver, spleen, erythron, internal bleeding in GI tracts, and other sources (Nairz et al. 2016; Weiss et al. 2019b).

Here, we demonstrate that EWAS is an effective computational approach to identify the association between a wide range of environmental exposures and multiple anemia phenotypes. Further research on the interactions between environmental risk exposures and genetic loci may provide an understanding of the mechanisms driving these complex phenotypes from multiple dimensions. Our findings not only demonstrate the power of evaluating environmental risk for anemias but also suggests that EWAS can reveal the environmental etiology of other complex diseases.

## Data Availability

For replication of our study, we provide links in the Supplementary file to all the raw data files, the R codes in R Markdown format (.Rmd), and corresponding reports (.nb.html), and all the results that were generated by using the R codes.

## Acknowledgment

The authors acknowledge the contribution made by Dr. Robert F. Paulson (The Pennsylvania State University) and Fenghua Qian (The Pennsylvania State University), who provided the knowledge on anemia; Anastasia M. Lucas (University of Pennsylvania), who established the *CLARITE* package and *hudson* package in R Program; and Lin Song (University of Wisconsin-Madison), who helped with proofreading and finalizing the manuscript. The authors sincerely acknowledge the financial support from the USDA National Institute of Food and Agriculture and Hatch Appropriations under Project #PEN04275 and Accession #1018544.

## Declaration of competing financial interests (CFI)

The authors declare they have no actual or potential competing financial interests.

