## Supplement Materials for "Environment-Wide Association Studies of Anemia in the National Health and Nutrition Examination Surveys"

<u>**Supplemental Material**</u>

### Abstract

*Background:*
Anemia is a global health problem that can lead to chronic illness in adults and may be fatal in children and the elderly. While some dietary factors and heavy metals are known risk factors for anemia, there are no environment-wide studies of anemia.

*Objectives:*
Our goal was to identify environment-wide risk factors for anemia.

*Methods:*
We evaluated general anemia in children and adults and further classified anemia as a) iron, vitamin B12, or folate deficiency anemia; b) anemia in general chronic diseases; and c) anemia in chronic kidney disease. As well as quantitative measures including level of hemoglobin, serum vitamin B12, red blood cell (RBC) folate, and serum iron. Environment-wide association studies (EWAS) were performed to identify novel environmental risk factors of anemia in discovery and replication subsets of the National Health and Nutrition Examination Surveys (NHANES).

*Results:*
We identified and replicated 106 potential environmental risk factors for anemia. As expected, serum iron was the top exposure associated with general anemia for adults. Cadmium was associated with adult hemoglobin levels, as were vitamin Bs, micronutrients, smoking, and alcohol consumption. Further, decreased levels of multiple vitamins, including vitamin A, vitamin Es and multiple vitamin Bs, were associated with general anemia in adults. Use of tobacco and alcohol was also found to be associated with red blood cell folate and serum iron levels. In children, serum iron level was associated with folic acid supplements and vitamin A supplements.

*Discussion:*
This is the first EWAS of anemia, providing insights into the environmental etiology of anemia risk in children and adults. These results may lead to the development of public health recommendations to mitigate anemia risk factors.

### Table of Contents

**Excel Table S1.** Variable dictionary for the post-QC variables with descriptions and categories. (**Will uploaded, right now could be accessed at** https://pennstateoffice365-my.sharepoint.com/:x:/g/personal/jpz5091_psu_edu/EQNV2epAZHBMuH8jHmeLNo4BrCN2Bfr1P-3Kg6nw-HRpqA?e=fZ3f02).

**Figure S1. Manhattan plot for general anemia EWAS results in children.** The statistical comparison was made between the anemic cohort and the non-risk control cohort with adjustment for age, sex, race, socioeconomic status, and survey year among children and teenagers (< 15 years old). The LRT was performed to compare the model with and without each categorical environmental factor by adjusting the same set of covariates. The x-axis shows the NHANES classified exposure categories, and the y-axis shows the -$\log_{10}$(p-value). The results from the discovery group were represented by circle and the results from the replication group were represented by triangle. The red line represents the FDR significant $p < 0.1$ in the discovery, and the blue line represents the Bonferroni significant $p < 0.05$ in the replication.

**Figure S2. Manhattan plot for general anemia EWAS results in adults** The statistical comparison was made between the anemic cohort and the non-risk control cohort by using binomial in GLM with adjustment for age, sex, race, socioeconomic status, survey year, and cotinine among young adults and adults (> 15 years old). The LRT was performed to compare the model with and without each categorical environmental factor by adjusting the same set of covariates. The x-axis shows the NHANES classified exposure categories, and the y-axis shows the -$\log_{10}$(p-value). The results from the discovery group were represented by circle and the results from the replication group were represented by triangle. The red line represents the FDR significant $p < 0.1$ in the discovery, and the blue line represents the Bonferroni significant $p < 0.05$ in the replication.

**Figure S3. Manhattan plot for EWAS results with serum hemoglobin in children.** The null hypothesis that the coefficient is equal to zero (no effect) was tested in a generalized linear model by adjusting for age, sex, race, socioeconomic status, survey year, cotinine, and iron supplement intake among children and teenagers (< 15 years old). The LRT was performed to compare the model with and without each categorical environmental factor by adjusting the same set of covariates. The x-axis shows the NHANES classified exposure categories, and the y-axis shows the -$\log_{10}$(p-value). The results from the discovery group were represented by circle and the results from the replication group were represented by triangle. The red line represents the FDR significant $p < 0.1$ in the discovery, and the blue line represents the Bonferroni significant $p < 0.05$ in the replication. p,p-DDT: clofenotane (dichlorodiphenyltrichloroethane). PCB: polychlorinated biphenyl.

**Figure S4. Manhattan plot for EWAS results with serum hemoglobin in adults.** The null hypothesis that the coefficient is equal to zero (no effect) was tested in a generalized linear model by adjusting for age, sex, race, socioeconomic status, survey year, cotinine, and iron supplement intake among young adults and adults (age larger than 15 years old). The LRT was performed to compare the model with and without each categorical environmental factor by adjusting the same set of covariates. The x-axis shows the NHANES classified exposure categories, and the y-axis shows the -$\log_{10}$(p-value). The results from the discovery group were represented by circle and the results from the replication group were represented by triangle. The red line represents the FDR significant $p < 0.1$ in the discovery, and the blue line represents the Bonferroni significant $p < 0.05$ in the replication. MFA 22:1: docosenoic.

**Figure S5. Manhattan plot for IDA EWAS results in female adults.** A statistical comparison was made between cases with IDA and the non-risk control cohort with adjustment for age, sex, race, socioeconomic status, survey year, cotinine, and pregnancy status for female young adults and adults (> 15 years old). The LRT was performed to compare the model with and without each categorical environmental factor by adjusting the same set of covariates. The x-axis shows the NHANES classified exposure categories, and the y-axis shows the -$\log_{10}$(p-value). The results from the discovery group were represented by circle and the results from the replication group were represented by triangle. The red line represents the FDR significant $p < 0.1$ in the discovery, and the blue line represents the Bonferroni significant $p < 0.05$ in the replication. RBC folate: folate level in red blood cells.

**Figure S6. Hudson plot comparing the EWAS results for blood iron concentrations in children comparing frozen serum samples (top) and combined frozen and refrigerated serum samples (bottom).** The null hypothesis that the coefficient is equal to zero (no effect) was tested in a generalized linear model by using the blood iron level from frozen serum (top) and combined measurements based on frozen serum and refrigerated serum (bottom) as the outcome and adjusting for age, sex, race, socioeconomic status, and survey year among children and teenagers (< 15 years old). The LRT was performed to compare the model with and without each categorical environmental factor by adjusting the same set of covariates. The red line represents the Bonferroni significant $p < 0.05$ in the discovery. The significant results were highlighted and labeled. All significant results are supplement uses except *LBXGTC* (serum γ-tocopherol) and *LBXVIA* (serum vitamin A, retinol).

**Figure S7. Manhattan plot for EWAS results with serum iron in children.** The null hypothesis that the coefficient is equal to zero (no effect) was tested in a generalized linear model by using the iron level from frozen serum as the outcome and adjusting for age, sex, race, socioeconomic status, and survey year among children and teenagers (< 15 years old). The LRT was performed to compare the model with and without each categorical environmental factor by adjusting the same set of covariates. The x-axis shows the NHANES classified exposure categories, and the y-axis shows the $-\log_{10}$(p-value). The results from the discovery group were represented by circle and the results from the replication group were represented by triangle. The red line represents the FDR significant $p < 0.1$ in the discovery, and the blue line represents the Bonferroni significant $p < 0.05$ in the replication.

**Figure S8. Manhattan plot for EWAS results with serum iron in adults.** The null hypothesis that the coefficient is equal to zero (no effect) was tested in a generalized linear model by using the iron level from frozen serum as the outcome and adjusting for age, sex, race, socioeconomic status, and survey year among young adults and adults (> 15 years old). The LRT was performed to compare the model with and without each categorical environmental factor by adjusting the same set of covariates. The x-axis shows the NHANES classified exposure categories, and the y-axis shows the $-\log_{10}$(p-value). The results from the discovery group were represented by circle and the results from the replication group were represented by triangle. The red line represents the FDR significant $p < 0.1$ in the discovery, and the blue line represents the Bonferroni significant $p < 0.05$ in the replication.

**Figure S9. Manhattan plot for CKD-A EWAS results in adults.** The statistical comparison was made between cases with CKD-A and the non-risk control cohort with adjustment for age, sex, race, socioeconomic status, survey year, and cotinine among young adults and adults (> 15 years old). The LRT was performed to compare the model with and without each categorical environmental factor by adjusting the same set of covariates. The x-axis shows the NHANES classified exposure categories, and the y-axis shows the $-\log_{10}$(p-value). The results from the discovery group were represented by circle. The red line represents the Bonferroni significant $p < 0.05$ in the discovery. RBC folate: folate level in red blood cells.

**Figure S10. Manhattan plot for UA EWAS results in children.** The statistical comparison was made between cases with UA and non-risk control cohort with adjustment for age, sex, race, socioeconomic status, survey year, and cotinine among children and teenagers (< 15 years old). The LRT was performed to compare the model with and without each categorical environmental factor by adjusting the same set of covariates. The x-axis shows the NHANES classified exposure categories, and the y-axis shows the $-\log_{10}$(p-value). The results from the discovery group were represented by circle. The red line represents the Bonferroni significant $p < 0.05$ in the discovery.

**Figure S11. Manhattan plot for UA EWAS results in adults.** The statistical comparison was made between cases with UA and the non-risk control cohort with adjustment for age, sex, race, socioeconomic status, survey year, and cotinine for young adults and adults (> 15 years old). The LRT was performed to compare the model with and without each categorical environmental factor by adjusting the same set of covariates. The x-axis shows the NHANES classified exposure categories, and the y-axis shows the $-\log_{10}$(p-value). The results from the discovery group were represented by circle and the results from the replication group were represented by triangle. The red line represents the FDR significant $p < 0.1$ in the discovery, and the blue line represents the Bonferroni significant $p < 0.05$ in the replication. RBC folate: folate level in red blood cells.

### External links

**Supplement Link S1.** All the raw data files could be accessed from shared folder at

https://pennstateoffice365-my.sharepoint.com/:f:/g/personal/jpz5091_psu_edu/Ej0HyvILa-FGspvskIdNU3cBPmjoLEITYfJiIUd1axT_Nw?e=cbHc7A.

File list:

| **File name** | **File type** | **Description** |
|---|---|---|
| MainTable.csv | File | The raw data table from (Patel et al. 2016). |
| MainTable_keepvar_over18.csv | File | The data table with the pre-selected variables and adults which was modified from MainTable.csv. |
| VarDescription.csv | File | The variable descriptions from (Patel et al. 2016). |
| VarCat_nopf.txt | File | The table with variables and their categories (groups). |
| data_adult.txt | File | The table for generating the Figure 2. |
| data_child.txt | File | The table for generating the Figure 3. |
| All_results_condensed_final.csv | File | The file from the R codes to create the network plot by using the Cytoscape (Shannon et al. 2003). |
| Data from NHANES website | Folder | This a folder that contains the raw data that were downloaded from the NHANES website: https://wwwn.cdc.gov/nchs/nhanes/Default.aspx |
| Weights | Folder | This a folder that contains the survey weights information from HNAES. |

**Supplement Link S2.** The R codes in R Markdown format (.Rmd) and corresponding report (.nb.html) could be accessed from

https://pennstateoffice365-my.sharepoint.com/:f:/g/personal/jpz5091_psu_edu/EoA6FFVeqK9CtULpeZdXclsBs8zp41ZeyKZY4tSLyPBHzg?e=XhqlPo.

File list:

| **File name** | **Description** |
|---|---|
| EWAS_Anemia_Notebook_EHP.Rmd | The R markdown file that contains all the codes. |
| EWAS_Anemia_Notebook_EHP.nb.html | A notebook in html file that contains all the codes and generated results from the R markdown file (EWAS_Anemia_Notebook_EHP.Rmd). |
| Code for classify Female and Ethnicity.rtf | R code for understanding the number of people in each ethnicity (Caucasian American, African American, Mexican American, other Hispanic, and other ethnicity) and sex (male and female). |

**Supplement Link S3.** All the results that were generated by using the R codes could be accessed from

https://pennstateoffice365-my.sharepoint.com/:f:/g/personal/jpz5091_psu_edu/EkbgXPfRbOFDti7KDvQVRKMBswJV78ouGnvXl5v4B-HWBA?e=o8RwUj.

File list:

| **File name** | **Description** | **Originated from** | **Related figures** |
|---|---|---|---|
| All_results_dis_adults.txt | All EWAS results from the discovery analysis for adults. | R codes | Figure 2 and Figure 4 |
| All_results_dis_children.txt | All EWAS results from the discovery analysis for children. | R codes | Figure 3 and Figure 5 |
| All_results_rep_adults.txt | All EWAS results from the replication analysis for adults. | R codes | Figure 2 and Figure 4 |
| All_results_rep_children.txt | All EWAS results from the replication analysis for children. | R codes | Figure 3 and Figure 5 |
| EWAS_[**Phenotype**]_[**Population**]_[**Setting**]_results.txt | EWAS results from the [**Setting**: discovery or replication] analysis for the [**Population**: Adult or Children] and [**Phenotype**: Anemia (Anemia), CKDA (Chronic Kidney Disease-related Anemia), hemo (hemoglobin), IDA (Iron Deficiency Anemia), IRN (Iron), RBF (Folate in Red Blood Cell), UA (Unexplained Anemia), and vb12 (serum Vitamin B12 concentration)]. | R codes | Figure 6 to Figure 20 |
| EWAS_[**Phenotype**]_[**Population**]_[**Setting**]_results_adj.txt | EWAS results from the [**Setting**: discovery or replication] analysis for the [**Population**: Adult or Children] and [**Phenotype**: Anemia (Anemia), CKDA (Chronic Kidney Disease-related Anemia), hemo (hemoglobin), IDA (Iron Deficiency Anemia), IRN (Iron), RBF (Folate in Red Blood Cell), UA (Unexplained Anemia), and vb12 (serum Vitamin B12 concentration)] with Bonferroni and FDR adjustments. | R codes | Figure 6 to Figure 20 |
